# Music therapy as a migraine treatment: protocol for a systematic review and meta-analysis

**DOI:** 10.1101/2021.02.24.21252365

**Authors:** Alessio Alberto Gilardi Sanchez, Milena Ana Micaela Guevara Bartolini, Maria Luz Pantoja Acosta, Maria Lazo-Porras

## Abstract

**Introduction:** Headache disorders are one of the most common health problems worldwide, they can be classified into primary and secondary disorders. In the primary group, migraine - the second most common type of headache - is the most disabling one and one of the most important reasons why its treatment is mandatory. Migraine treatment involves different steps and kinds of medical therapy and patient education, and in these past years studies have been exploring the effect music therapy can have in reducing the severity and duration of an acute migraine attack. It has been reported that adding music to the pharmacological treatment can help decrease the pain severity, thus, reducing the disability migraine can cause.

**Objective:** Evaluate the effectiveness of music therapy as a treatment or coadjuvant of migraine attacks in people who suffer this condition.

**Methods and analysis:** This protocol is consistent with the methodology recommended by the PRISMA-P and the Cochrane handbook for systematic reviews of interventions. This study will be carried out as a systematic review and meta-analysis. In order to do so, electronic searches will be performed in PubMed, Medline and Cochrane (through Ovid) and Embase. The data range parameters used in searching all databases are from the last 20 years. Randomized controlled trials (RCTs) published in English, Spanish, French and Portuguese; with the primary outcomes being reduction of headache intensity, resolution of the migraine and decreased frequency of migraine attacks. Three investigators will screen all retrieved studies titles and abstracts, making a first preliminary list. A second screen will be done by the same three investigators similarly to the first one, but reviewing the full texts and building the final list. Then, the evaluation of the risk of bias and extraction of all data will be performed. The risk of bias of the included RCTs will be evaluated by the Cochrane Collaboration’s tool. A qualitative synthesis will be provided in text and tables, to summarize the main results of the selected publications. The heterogeneity between studies will be assessed through the *I*^*2*^ statistic. If there is sufficient homogeneity across outcomes, a meta-analysis will be conducted.

**Conclusions:** This systematic review will provide evidence regarding the effectiveness of music therapy as a single or coadjuvant treatment in patients with migraine attacks. Based on this analysis, it will be feasible to know whether this intervention is effective in the reduction of the intensity of the migraine attack, if it can help resolve the migraine attack, or reduce the frequency of migraine attacks.

## INTRODUCTION

Headache disorders are one of the most common health problems worldwide. They are thought to be the fifth cause of disability-adjusted life-years in people between 25 and 49 years old ^1^. Headache disorders can be classified mainly into primary and secondary disorders. Primary headache disorders include migraine, tension-type headache, trigeminal autonomic cephalalgias and other less common types like primary cough headache, primary exercise headache, etc ^2,3^. They are called primary headache disorders because they are thought to be caused by intrinsic cerebral causes and not by an identifiable pathology or disease ^3^, as the secondary headache disorders. The latter include headaches attributed to traumas, infections, vascular disorders, psychiatric disorders, among others ^2^.

Tension-type headache is the most common primary headache disorder, having a prevalence of approximately 38.3%, followed by migraine with approximately 12% in the US but 9% in South America, and lastly the trigeminal autonomic cephalgias with a prevalence of approximately 0.1% in the general population ^4^. Although tension-type headache is more common than migraine, it only causes 7.2 million years of life lived with disability (YLDs) globally each year, which is a small proportion compared to the 45.1 million YLDs that migraine causes ^5^. In 2001, a survey made in the US showed that migraine affects about 12% of the population, being predominant in women over men: 18.2% and 6.5%, respectively ^6^. The same predominance has been reported in China, Japan, and South Korea, where prevalence of migraine among adults ranged from 6% to 14.3% (11-20% among women and 3-8% among men) ^7^, similarly to what’s seen in the US. In Peru, a study conducted in Cuzco in 1997 showed a lower migraine prevalence: 5.3%, although, predominance among women over men can be seen in this population too (2.3% vs 7.8%) ^8^.

Even though migraine accounts for more disability than all the other neurologic disorders combined ^5^, its pathophysiology is not fully yet understood. The most accepted theory is that the trigeminovascular system is the anatomical origin of migraine. From there, the first-order neurons innervate the meninges and their vessels and the second-order neurons in the brain stem. From there, the second-order neurons activate the third-order neurons located in the thalamus, which innervate the cortex and create the perception of pain ^9^. It is thought that activation of the trigeminovascular system causes an outflow to the intracranial arteries, which causes vasodilation due to molecules such as pituitary adenylate cyclase–activating peptide (PACAP) and, in turn, pain. The reason why the trigeminovascular system is originally activated is not known yet ^10^.

Migraine can be subclassified into different clinical presentations like migraine with aura– which can be subclassified into more specific pathologies–, without aura and chronic migraine ^2^. Each kind of migraine has its own characteristics and diagnostic criteria. For example, migraine without aura is characterized by at least 5 episodes of headache attacks lasting 4-72 hours; at least 2 characteristics between: unilateral location, pulsating quality, moderate or severe intensity, aggravation by or causing avoidance of physical activity; and at least one of the following: nausea and/or vomiting, photophobia and phonophobia ^2^. Even though migraine with and without aura are distinguishable diagnostics, they don’t rule out each other, meaning that a person who suffers from migraine without aura can have attacks with aura ^11^. Furthermore, migraine without aura attacks can have prodromal symptoms, which may be present hours or days before the attacks, originate in different parts of the brain, as opposed to aura, which is involved in a specific cortical area ^2^.

Being migraine such a common and disabling condition, its treatment and prevention is of utmost importance ^5^. Migraine treatment involves different steps and kinds of treatment, depending on when and where it is being administered, the severity of the attack, and its recurrence. Migraine treatment first steps, once the diagnosis has been established, involve educating the patient about the pathology; establishing the patient’s expectations about the disease and its treatment; encouraging the patient to identify triggers; among others ^12^.

There are some studies that suggest that music therapy may help alleviate pain such as cancer pain ^13^, procedural pain ^14^ and it may have a great effect in reducing the severity and duration of an acute migraine attack ^15^. It has been reported that adding music to the pharmacological treatment helps decrease the pain severity, as pain impulses travel from the site of injury to the brain throughout the spinal cord in which neural gates indicate the level of pain that is allowed to pass or not to pass. As a patient listens to music, the gate closes resulting in a decrease of the pain impulses that reach the brain. The brain itself releases neurotransmitters as an answer to music which stimulates the release of endorphins, these modulate pain and as an effect is reflected a feeling of euphoria. In conclusion, listening to music by inhibiting pain and stimulating the release of endorphins, the patient’s migraine improves ^15^.

### Justification

Migraine is one of the most common causes of headache worldwide. As usually known, it is a primary headache whose main features are recurrent unilateral headache with localized pain; usually accompanied with nausea and/or vomiting; intolerance of light and or sound (such as extremely loud and prolonged noises or music such as concerts, fireworks, constructions); and that may be exacerbated by physical activity ^2^. In 25% of people who suffer this condition, it is accompanied with visual, sensory and or speech aura preceding the migraine attack that lasts less than 60 minutes. As it is a disabling condition most of the time, it is fundamental to know the triggers that cause the attack and have an abortive therapy depending on the type of migraine. If it is a mild to moderate migraine, the first line treatment is usually acetaminophen, NSAIDs and it may be combined with caffeine. A specific migraine agent such as triptans or ergotamine are indicated in a moderate to severe migraine. Besides the pharmacological treatment, it is important to make some lifestyle modifications such as a regular sleeping schedule, exercise or diet to avoid triggers that could perpetuate the migraine attack, and other kinds of treatments. Among these, in the majority of studies, listening to music while having the episode and in combination with medical therapy, has shown an improvement in decreasing the headache pain and helping to return to the patient’s basal state before the migraine ^16^. This meta-analysis is about evaluating the effectiveness of musical therapy in the acute migraine attack.

### Objective

To evaluate the effectiveness of music therapy as a treatment or coadjuvant of migraine attacks in people who suffer this condition.

## METHODS AND ANALYSES

### Study design

This study will consist of a systematic review and meta-analysis, which will be conducted and published according to the Preferred Reporting Items for Systematic Reviews and Meta-Analysis Protocols (PRISMA-P) ^17,18^.

### Eligibility criteria

#### Types of studies

This Meta-Analysis will include all randomized controlled trials (RCT) published in Spanish, English, French or Portuguese that investigated the effectiveness of music therapy in the treatment of migraine in people with this condition. The publication date restriction of the individual studies will be from 2000 and later.

#### Participants

This study will include all RCTs that recruited children, adolescents and/or adult patients with diagnosis of migraine with aura, without aura or chronic migraine according to the 2nd or 3rd edition of The International Classification of Headache Disorders ^2,19^. Patients with severe migraine will be included, however, multiple scales and tools for assessing migraine severity ^20–24^ exist and some are still under development ^25^. No tool has still been recognized as the gold standard for severity classification, patients with any kind of severity assigned by any scale used in each individual study, if done, will be considered eligible.

#### Intervention

Trials that evaluate music therapy as a single treatment of acute migraine attacks and/or as a coadjuvant to a single or double pharmacological therapy. Interventions could have been performed individually or in groups; in a care centre or at home; supervised by therapists or self-assessed; and with a single or multiple genres of music. No limits will be imposed on the timing, frequency and duration of the interventions.

#### Comparison

Any control intervention like no treatment, pharmacological treatment (single or double therapy) or placebo.

#### Types of outcome measures

Primary outcomes

- Reduction in headache intensity
- Resolution of headache
- Decreased frequency of migraine attacks

Secondary outcomes

- Resume of routine activities
- Associated symptoms (e.g. nausea or vomiting)
- Duration of migraine attacks (hours)
- Number of migraine episodes with severe intensity
- Impact of migraine on everyday level of functioning

### Exclusion criteria

- Any of the following study designs: cross-sectional, case control, uncontrolled, cohort, non-randomized trials, pilots, case reports, case series, letter to the editor, editorial, observational and epidemiological studies, correspondence, narrative review and systematic reviews (with or without meta-analysis).
- Studies that evaluate music therapy for status migrainosus.
- Studies that evaluate music therapy as a coadjuvant of non-pharmacological therapies (diet, acupuncture, etc.)
- Studies that compare music therapy to other kind of treatment other than placebo, no treatment or pharmacological treatments
- Studies published in any languages other than Spanish, English, Portuguese or French.
- Studies that are not accessible in full text despite being requested to the author.

### Information sources

Electronic searches will be performed for potentially eligible RCTs in PubMed, Medline and Cochrane (through Ovid), and Embase with restriction in articles with full texts in English, Spanish, French and Portuguese published since 2000.

Different combinations of the following keywords in English will be used: migraine, migrainous disorder, migraine attack, musical, music, treatment, therapy, therapeutic, randomized controlled trial, clinical trial, among others, to search for the most data available.

### Search strategy

In each database, different combinations of the keywords previously mentioned will be developed to maximize the search and try to find all the studies that meet the eligibility criteria for this meta-analysis. The preliminary search strategies are shown in the **Appendix 1** for each of the four databases used.

#### Study records

##### Data management

The software that will be used to help upload, store and select the results will be *Rayyan QCRI*. Each database will have a separate library group that will be created to keep the original search results. Then, all separate library group copies will be merged into a new library group and duplicate checking will be carried out in the new library group.

##### Selection process

Once the search has been completed and the studies have been stored, the primary selection will be done by three different investigators. They will select, independently, all the studies that meet the inclusion criteria based on the titles and abstracts. Once each investigator has completed the selection process, the literature selected by each investigator will be compared with the results selected by the other two and the differences will be discussed and resolved, deciding if the studies should be included or not. After the first preliminary selection has been done, the same three investigators will review the complete studies and select the ones that fulfill the inclusion criteria. Once the secondary selection has been completed, the three lists will be compared and the differences will be reviewed and discussed, as in the first step. Once this discussion is done, the final list of studies will be completed. These studies will be the ones that will be included, discussed and analyzed.

A Preferred Reporting Items for Systematic Reviews and Meta-Analysis (PRISMA) flow diagram will be done to show each phase’s number of included and excluded studies.

##### Data collection process

Two independent authors will carry out the data extraction. The extracted data will include general study information, characteristics of participants, characteristics of interventions, characteristics of the comparison treatment and outcome measures. If necessary, the corresponding authors of the selected publications will be contacted for missing data and further information. Disagreements between the two reviewers will be solved by a third reviewer to achieve a consensus.

Three authors will record the results of included and excluded studies and show them in a PRISMA flow diagram.

#### Data items

##### The extracted data will include

- Study details: First author, corresponsal author, article title, multicenter characteristics, country/countries, publication year, language, randomized controlled trial.
- Participant’s characteristics: Sample size (number of participants in each group/arm), inclusion and exclusion criteria, age range, gender, randomization process, type of migraine (mild/moderate/severe), migraine details (with/without aura)
- Details of interventions: type of intervention, duration, frequency, type of music, in which moment is music therapy indicated to be used, supervision, implementer, arm segment, control or comparison group, etc.
- Outcome measures: severity of the migraine, resolution of the migraine attack. The secondary outcomes mentioned in the section “Types of outcome measures” will also be collected.

If data from a study is as reported in more than one article, it will only be selected from the article with the most complete data for this research purposes.

### Risk of bias in individual studies

The risk of bias will be managed using the Cochrane Bias Tool for RCT, which assesses the following domains in the measurement of results and in the selective reporting of results: biases in the randomization process, due to deviations from the planned interventions, due to incomplete results data. The classification of the biases will be low risk, some concerns and high risk ^26^.

These risks of bias judgments will be made by three independent reviewers and disagreements will be resolved discussing them.

### Analysis

In this study, continuous and dichotomous effect sizes will be synthesized and a meta-analysis with a random-effects and intention-to-treat approach will be performed. For each outcome, the between-study variance will be estimated with the Restricted Maximum Likelihood (REML). Additionally, the method to calculate the uncertainty in the overall effect size, namely the 95% confidence interval (CI), will be with the Hartung-Knapp-Sidik-Jonkman (HKSJ) method.

Each study will report effect sizes or aggregated raw data (total events and participants or mean and standard deviation). When both are reported, only the effect size, if adjusted, will be used. For continuous outcomes, the mean differences (MD) with their corresponding 95% CI for outcomes that use the same scale, questionnaire or score will be used or calculated. If different instruments were used, the standardized mean differences (SMD) and their 95% CI will be calculated. For all these calculations, the mean and standard deviation reported after the intervention and not the change in means will be used. Similarly, for dichotomous outcomes, the risk ratios (RR) with their 95% CI will be either used or calculated.

Heterogeneity across studies will be assessed with the *I*^*2*^ and visually. Roughly, an *I*^*2*^ <25% may reflect low inconsistency and *I*^*2*^ >50% high inconsistency. Results with high inconsistency will be explored and sub-analysis will be performed based on their differences in population, intervention, comparison and outcome. Subgroup analysis will not be performed. The analysis will be made with RevMan 5.4.1.

## Data Availability

All relevant information will be published as supplementary materials.

## ETHICS AND DISSEMINATION

Approval of the project will be requested to the Ethics Committee of the “Universidad Peruana Cayetano Heredia”. The results of this review will be disseminated through peer-reviewed publications.

### APPENDIX

#### Appendix 1. Preliminary search strategies

##### A.Pubmed

###### Descriptor 1: Migraine

120,653 results

###### MeSH terms

“Migraine Disorders”[Mesh] OR “Migraine without Aura”[Mesh] OR “Migraine with Aura”[Mesh] OR “Headache”[Mesh] OR “Headache Disorders, Primary”[Mesh] OR “Headache Disorders”[Mesh]

###### Free terms

“Migraine Disorder” OR “Migraine Disorders” OR “Migraine without Aura” OR “Migraine with Aura” OR “Migraine” OR “Migraine Attack” OR “Migraine Attacks” OR “Migrainous” OR “Migrainous Disorder” OR “Migrainous Disorders” OR “Migraines” OR “Acute Migraine” OR “Chronic Migraine” OR “Headache” OR “Headaches” OR “Headache Disorders” OR “Primary Headache Disorders”

###### MeSH terms + Free terms

“Migraine Disorders”[Mesh] OR “Migraine without Aura”[Mesh] OR “Migraine with Aura”[Mesh] OR “Headache”[Mesh] OR “Headache Disorders, Primary”[Mesh] OR “Headache Disorders”[Mesh] OR “Migraine Disorder” OR “Migraine Disorders” OR “Migraine without Aura” OR “Migraine with Aura” OR “Migraine” OR “Migraine Attack” OR “Migraine Attacks” OR “Migrainous” OR “Migrainous Disorder” OR “Migrainous Disorders” OR “Migraines” OR “Acute Migraine” OR “Chronic Migraine” OR “Headache” OR “Headaches” OR “Headache Disorders” OR “Primary Headache Disorders”

###### Descriptor 2: Music therapy

446,172 results

###### MeSH terms

“Music Therapy”[Mesh] OR “Psychotherapy”[Mesh] OR “Sensory Art Therapies”[Mesh] OR “Complementary Therapies”[Mesh] OR “Music”[Mesh]

###### Free terms

“Music Therapy” OR “Music” OR “Sensory Art Therapies” OR “Psychotherapy” OR “Musical” OR “Musicotherapy” OR “Complementary Therapies” OR “Music Therapies” OR “Complementary Therapy” OR “Sensory Art Therapy”

###### MeSH terms + Free terms

“Music Therapy”[Mesh] OR “Psychotherapy”[Mesh] OR “Sensory Art Therapies”[Mesh] OR “Complementary Therapies”[Mesh] OR “Music”[Mesh] OR “Music Therapy” OR “Music” OR “Sensory Art Therapies” OR “Psychotherapy” OR “Musical” OR “Musical” OR “Musicotherapy” OR “Complementary Therapies” OR “Music Therapies” OR “Complementary Therapy”

###### Descriptor 3: Randomized controlled trials (RCTs)

4,405,384 results

###### MeSH terms

“Randomized Controlled Trial” [Publication Type] OR “Randomized Controlled Trials as Topic”[Mesh] OR “Controlled Clinical Trial” [Publication Type]

###### Free terms

“Randomized Controlled Trial” OR “Randomized Controlled Trials” OR “Randomized Trial” OR “Randomized Trials” OR “Controlled Clinical Trial” OR “Controlled Clinical Trials” OR “RCT” OR “RCTs” OR “Controlled Trial” OR “Controlled trials” OR (randomized controlled trial[pt] OR controlled clinical trial[pt] OR randomized[tiab] OR placebo[tiab] OR drug therapy[sh] OR randomly[tiab] OR trial[tiab] OR groups[tiab] NOT (animals [mh] NOT humans [mh]))

###### B.MeSH terms + Free terms

“Randomized Controlled Trial” [Publication Type] OR “Randomized Controlled Trials as Topic”[Mesh] OR “Controlled Clinical Trial” [Publication Type] OR “Randomized Controlled Trial” OR “Randomized Controlled Trials” OR “Randomized Trial” OR “Randomized Trials” OR “Controlled Clinical Trial” OR “Controlled Clinical Trials” OR “RCT” OR “RCTs” OR “Controlled Trial” OR “Controlled trials” OR (randomized controlled trial[pt] OR controlled clinical trial[pt] OR randomized[tiab] OR placebo[tiab] OR drug therapy[sh] OR randomly[tiab] OR trial[tiab] OR groups[tiab] NOT (animals [mh] NOT humans [mh]))

###### All (MeSH terms + Free terms)

Filters applied: English, French, Portuguese, Spanish and studies developed from the year 2000 forward 1,170 results

(“Migraine Disorders”[Mesh] OR “Migraine without Aura”[Mesh] OR “Migraine with Aura”[Mesh] OR “Headache”[Mesh] OR “Headache Disorders, Primary”[Mesh] OR “Headache Disorders”[Mesh] OR “Migraine Disorder” OR “Migraine Disorders” OR “Migraine without Aura” OR “Migraine with Aura” OR “Migraine” OR “Migraine Attack” OR “Migraine Attacks” OR “Migrainous” OR “Migrainous Disorder” OR “Migrainous Disorders” OR “Migraines” OR “Acute Migraine” OR “Chronic Migraine” OR “Headache” OR “Headaches” OR “Headache Disorders” OR “Primary Headache Disorders”) AND (“Music Therapy”[Mesh] OR “Psychotherapy”[Mesh] OR “Sensory Art Therapies”[Mesh] OR “Complementary Therapies”[Mesh] OR “Music”[Mesh] OR “Music Therapy” OR “Music” OR “Sensory Art Therapies” OR “Psychotherapy” OR “Musical” OR “Musical” OR “Musicotherapy” OR “Complementary Therapies” OR “Music Therapies” OR “Complementary Therapy”) AND (“Randomized Controlled Trial” [Publication Type] OR “Randomized Controlled Trials as Topic”[Mesh] OR “Controlled Clinical Trial” [Publication Type] OR “Randomized Controlled Trial” OR “Randomized Controlled Trials” OR “Randomized Trial” OR “Randomized Trials” OR “Controlled Clinical Trial” OR “Controlled Clinical Trials” OR “RCT” OR “RCTs” OR “Controlled Trial” OR “Controlled trials” OR (randomized controlled trial[pt] OR controlled clinical trial[pt] OR randomized[tiab] OR placebo[tiab] OR drug therapy[sh] OR randomly[tiab] OR trial[tiab] OR groups[tiab] NOT (animals [mh] NOT humans [mh])))

##### B.Medline and Cochrane (through Ovid) Descriptor 1: Migraine

156,300 results

###### Indexed terms

exp migraine disorders/ or exp headache disorders, primary/ or exp headache disorders/ or exp migraine with aura/ or exp migraine without aura/ or exp headache

###### Free terms

(migraine$ or migraine with aura$ or migraine without aura$ or migraine disorder$ or migraine attack$ or migrainous$ or acute migraine$ or chronic migraine$ or headache$ or headache disorder$ or primary headache disorder$).mp.

###### Indexed terms + Free terms

exp migraine disorders/ or exp headache disorders, primary/ or exp headache disorders/ or exp migraine with aura/ or exp migraine without aura/ or exp headache/ or (migraine$ or migraine with aura$ or migraine without aura$ or migraine disorder$ or migraine attack$ or migrainous$ or acute migraine$ or chronic migraine$ or headache$ or headache disorder$ or primary headache disorder$).mp.

###### Descriptor 2: Music therapy

465,156 results

###### Indexed terms

exp music/ or exp music therapy/ or exp sensory art therapy/ or exp psychotherapy/ or exp complementary therapies

###### Free terms

(music therapy$ or music$ or sensory art therapy$ or psychotherapy$ or musical$ or musicotherapy$ or complementary therapy$ or music therapy$).mp.

###### Indexed terms + Free terms

exp music/ or exp music therapy/ or exp sensory art therapy/ or exp psychotherapy/ or exp complementary therapies/ or (music therapy$ or music$ or psychotherapy$ or musical$ or musicotherapy$ or complementary therapy$ or music therapy$).mp.

###### Descriptor 3: Randomized controlled trials (RCTs)

1,844,297 results

###### Indexed terms

exp randomized controlled trial/ or exp controlled clinical trial/ or exp randomized controlled trials as topic/ or exp clinical trials as topic

###### Free terms

(randomized controlled trial$ or controlled trial$ or randomized trial$ or controlled clinical trial$ or rct$).mp.

###### Indexed terms + Free terms

exp randomized controlled trial/ or exp controlled clinical trial/ or exp randomized controlled trials as topic/ or exp clinical trials as topic/ or (randomized controlled trial$ or controlled trial$ or randomized trial$ or controlled clinical trial$ or rct$).mp.

###### All (Indexed terms + Free terms)

Filters applied: English, French, Portuguese, Spanish and studies developed from the year 2000 forward

1,159 results (exp migraine disorders/ or exp headache disorders, primary/ or exp headache disorders/ or exp migraine with aura/ or exp migraine without aura/ or exp headache/ or (migraine$ or migraine with aura$ or migraine without aura$ or migraine disorder$ or migraine attack$ or migrainous$ or acute migraine$ or chronic migraine$ or headache$ or headache disorder$ or primary headache disorder$).mp.) and (exp music/ or exp music therapy/ or exp sensory art therapy/ or exp psychotherapy/ or exp complementary therapies/ or (music therapy$ or music$ or psychotherapy$ or musical$ or musicotherapy$ or complementary therapy$ or music therapy$).mp.) and (exp randomized controlled trial/ or exp controlled clinical trial/ or exp randomized controlled trials as topic/ or exp clinical trials as topic/ or (randomized controlled trial$ or controlled trial$ or randomized trial$ or controlled clinical trial$ or rct$).mp.)

##### C.Embase

###### Descriptor 1: Migraine

307,200 results

###### Emtree Terms

(’migraine’/exp OR ’migraine’ OR ’migraine with aura’/exp OR ’migraine with aura’ OR ’migraine without aura’/exp OR ’migraine without aura’ OR ’primary headache’/exp OR ’primary headache’ OR ’headache’/exp OR ’headache’ OR ’status migrainosus’/exp OR ’status migrainosus’)

###### Free Terms

(’migraine disorder’:ab,ti OR ’migraine disorders’:ab,ti OR ’migraine without aura’:ab,ti OR ’migraine with aura’:ab,ti OR ’migraine’:ab,ti OR ’migraine attack’:ab,ti OR ’migraine attacks’:ab,ti OR ’migrainous’:ab,ti OR ’migrainous disorder’:ab,ti OR ’migrainous disorders’:ab,ti OR ’migraines’:ab,ti OR ’acute migraine’:ab,ti OR ’chronic migraine’:ab,ti OR ’headache’:ab,ti OR ’headaches’:ab,ti OR ’headache disorders’:ab,ti OR ’primary headache disorders’:ab,ti)

###### Emtree Terms + Free Terms

(’migraine’/exp OR ’migraine’ OR ’migraine with aura’/exp OR ’migraine with aura’ OR ’migraine without aura’/exp OR ’migraine without aura’ OR ’primary headache’/exp OR ’primary headache’ OR ’headache’/exp OR ’headache’ OR ’status migrainosus’/exp OR ’status migrainosus’) OR (’migraine disorder’:ab,ti OR ’migraine disorders’:ab,ti OR ’migraine without aura’:ab,ti OR ’migraine with aura’:ab,ti OR ’migraine’:ab,ti OR ’migraine attack’:ab,ti OR ’migraine attacks’:ab,ti OR ’migrainous’:ab,ti OR ’migrainous disorder’:ab,ti OR ’migrainous disorders’:ab,ti OR ’migraines’:ab,ti OR ’acute migraine’:ab,ti OR ’chronic migraine’:ab,ti OR ’headache’:ab,ti OR ’headaches’:ab,ti OR ’headache disorders’:ab,ti OR ’primary headache disorders’:ab,ti)

###### Descriptor 2: Music therapy

69,695 results

###### Emtree Terms

(’music therapy’/exp OR ’music therapy’ OR ’music therapeutic use’/exp OR ’music therapeutic use’ OR ’music therapist’/exp OR ’music therapist’)

###### Free Terms

(’music therapy’:ab,ti OR ’music’:ab,ti OR ’sensory art therapies’:ab,ti OR ’psychotherapy’:ab,ti OR ’musical’:ab,ti OR ’musicotherapy’:ab,ti OR ’complementary therapies’:ab,ti OR ’music therapies’:ab,ti OR ’complementary therapy’:ab,ti OR ’sensory art therapy’:ab,ti)

###### Emtree Terms + Free Terms

(’music therapy’/exp OR ’music therapy’ OR ’music therapeutic use’/exp OR ’music therapeutic use’ OR ’music therapist’/exp OR ’music therapist’) OR (’music therapy’:ab,ti OR ’music’:ab,ti OR ’sensory art therapies’:ab,ti OR ’psychotherapy’:ab,ti OR ’musical’:ab,ti OR ’musicotherapy’:ab,ti OR ’complementary therapies’:ab,ti OR ’music therapies’:ab,ti OR ’complementary therapy’:ab,ti OR ’sensory art therapy’:ab,ti)

###### Descriptor 3: Randomized controlled trials (RCTs)

2,580,871 results

###### Emtree terms

(’randomized controlled trial’/exp OR ’randomized controlled trial’ OR ’randomized controlled trial topic’/exp OR ’randomized controlled trial topic’ OR ’controlled clinical trial’/exp OR ’controlled clinical trial’)

###### Free terms

(’randomised controlled study’:ab,ti OR ’randomised controlled trial’:ab,ti OR ’rct’:ab,ti OR ’non-rct’:ab,ti OR ’comparative studies’:ab,ti OR ’control group study’:ab,ti OR ’control group trial’:ab,ti OR ’controlled trial’:ab,ti OR ’experimental studies’:ab,ti OR ’feasibility studies’:ab,ti OR ’field studies’:ab,ti OR ’non experimental studies’:ab,ti OR ’non experimental study’:ab,ti OR ’nonexperimental studies’:ab,ti OR ’nonexperimental study’:ab,ti OR ’observation studies’:ab,ti OR ’observation study’:ab,ti OR ’observational studies’:ab,ti OR ’panel study’:ab,ti OR ’prevention trial’:ab,ti OR ’preventive study’:ab,ti OR ’preventive trial’:ab,ti OR ’quality improvement studies’:ab,ti OR ’quasiexperimental study’:ab,ti OR ’replication studies’:ab,ti OR ’trend studies’:ab,ti OR ’twin studies’:ab,ti OR ’twins study’:ab,ti OR ’validation studies’:ab,ti OR random*:ab,ti OR factorial*:ab,ti OR crossover*:ab,ti OR ((cross NEXT/1 over*):ab,ti) OR placebo*:ab,ti OR (doubl*:ab,ti AND blind*:ab,ti) OR (singl*:ab,ti AND blind*:ab,ti) OR assign*:ab,ti OR volunteer*:ab,ti)

###### Emtree Terms + Free Terms

(’randomized controlled trial’/exp OR ’randomized controlled trial’ OR ’randomized controlled trial topic’/exp OR ’randomized controlled trial topic’ OR ’controlled clinical trial’/exp OR ’controlled clinical trial’) OR (’randomised controlled study’:ab,ti OR ’randomised controlled trial’:ab,ti OR ’rct’:ab,ti OR ’non-rct’:ab,ti OR ’comparative studies’:ab,ti OR ’control group study’:ab,ti OR ’control group trial’:ab,ti OR ’controlled trial’:ab,ti OR ’experimental studies’:ab,ti OR ’feasibility studies’:ab,ti OR ’field studies’:ab,ti OR ’non experimental studies’:ab,ti OR ’non experimental study’:ab,ti OR ’nonexperimental studies’:ab,ti OR ’nonexperimental study’:ab,ti OR ’observation studies’:ab,ti OR ’observation study’:ab,ti OR ’observational studies’:ab,ti OR ’panel study’:ab,ti OR ’prevention trial’:ab,ti OR ’preventive study’:ab,ti OR ’preventive trial’:ab,ti OR ’quality improvement studies’:ab,ti OR ’quasiexperimental study’:ab,ti OR ’replication studies’:ab,ti OR ’trend studies’:ab,ti OR ’twin studies’:ab,ti OR ’twins study’:ab,ti OR ’validation studies’:ab,ti OR random*:ab,ti OR factorial*:ab,ti OR crossover*:ab,ti OR ((cross NEXT/1 over*):ab,ti) OR placebo*:ab,ti OR (doubl*:ab,ti AND blind*:ab,ti) OR (singl*:ab,ti AND blind*:ab,ti) OR assign*:ab,ti OR volunteer*:ab,ti)

###### All terms (Emtree Terms + Free Terms)

Filters applied: English, French, Portuguese, Spanish and studies developed from the year 2000 forward

244 results

(’migraine’/exp OR ’migraine’ OR ’migraine with aura’/exp OR ’migraine with aura’ OR ’migraine without aura’/exp OR ’migraine without aura’ OR ’primary headache’/exp OR ’primary headache’ OR ’headache’/exp OR ’headache’ OR ’status migrainosus’/exp OR ’status migrainosus’) OR (’migraine disorder’:ab,ti OR ’migraine disorders’:ab,ti OR ’migraine without aura’:ab,ti OR ’migraine with aura’:ab,ti OR ’migraine’:ab,ti OR ’migraine attack’:ab,ti OR ’migraine attacks’:ab,ti OR ’migrainous’:ab,ti OR ’migrainous disorder’:ab,ti OR ’migrainous disorders’:ab,ti OR ’migraines’:ab,ti OR ’acute migraine’:ab,ti OR ’chronic migraine’:ab,ti OR ’headache’:ab,ti OR ’headaches’:ab,ti OR ’headache disorders’:ab,ti OR ’primary headache disorders’:ab,ti) AND (’music therapy’/exp OR ’music therapy’ OR ’music therapeutic use’/exp OR ’music therapeutic use’ OR ’music therapist’/exp OR ’music therapist’) OR (’music therapy’:ab,ti OR ’music’:ab,ti OR ’sensory art therapies’:ab,ti OR ’psychotherapy’:ab,ti OR ’musical’:ab,ti OR ’musicotherapy’:ab,ti OR ’complementary therapies’:ab,ti OR ’music therapies’:ab,ti OR ’complementary therapy’:ab,ti OR ’sensory art therapy’:ab,ti) AND (’randomized controlled trial’/exp OR ’randomized controlled trial’ OR ’randomized controlled trial topic’/exp OR ’randomized controlled trial topic’ OR ’controlled clinical trial’/exp OR ’controlled clinical trial’) OR (’randomised controlled study’:ab,ti OR ’randomised controlled trial’:ab,ti OR ’rct’:ab,ti OR ’non-rct’:ab,ti OR ’comparative studies’:ab,ti OR ’control group study’:ab,ti OR ’control group trial’:ab,ti OR ’controlled trial’:ab,ti OR ’experimental studies’:ab,ti OR ’feasibility studies’:ab,ti OR ’field studies’:ab,ti OR ’non experimental studies’:ab,ti OR ’non experimental study’:ab,ti OR ’nonexperimental studies’:ab,ti OR ’nonexperimental study’:ab,ti OR ’observation studies’:ab,ti OR ’observation study’:ab,ti OR ’observational studies’:ab,ti OR ’panel study’:ab,ti OR ’prevention trial’:ab,ti OR ’preventive study’:ab,ti OR ’preventive trial’:ab,ti OR ’quality improvement studies’:ab,ti OR ’quasiexperimental study’:ab,ti OR ’replication studies’:ab,ti OR ’trend studies’:ab,ti OR ’twin studies’:ab,ti OR ’twins study’:ab,ti OR ’validation studies’:ab,ti OR random*:ab,ti OR factorial*:ab,ti OR crossover*:ab,ti OR ((cross NEXT/1 over*):ab,ti) OR placebo*:ab,ti OR (doubl*:ab,ti AND blind*:ab,ti) OR (singl*:ab,ti AND blind*:ab,ti) OR assign*:ab,ti OR volunteer*:ab,ti)

